# Novel deterministic epidemic model considering mass vaccination and lockdown against coronavirus disease 2019 spread in Israel: A numerical study

**DOI:** 10.1101/2021.05.15.21257264

**Authors:** Motoaki Utamura, Makoto Koizumi, Seiichi Kirikami

**Affiliations:** Research Laboratory for Nuclear Reactors, Tokyo Institute of Technology, 2-5-2-612 Kamiyoga Setagaya-ku Tokyo 158-0098 Japan; Hitachi Research Laboratory, Hitachi Ltd., Chiba city; Hitachi Works, Hitachi Ltd., Hitachi City

**Keywords:** COVID-19, Israel, vaccination, ATLM, epidemic model, discrete delay differential equation

## Abstract

Why public health intervention by the Israeli government against coronavirus disease 2019 spread has been successful while the majority of other countries are still coping with it? To give a quantitative answer, a simple numerical epidemic model is prepared to simulate the entire trend of various infection-related variables considering the 1^st^ and 2^nd^ vaccination campaigns against the alpha variant and simultaneous lockdown. This model is an extension of our previously published deterministic physical model, i.e. Apparent Time Lag Model, which aims at predicting an entire trend of variables in a single epidemic. The time series data of both vaccine dose ratio and lockdown period are employed in the model. Predictions have been compared with observed data in terms of daily new cases, isolated people, infections at large and effective reproductive number, and, further, the model is verified. Moreover, parameter survey calculations for several scenarios have clarified the synergy effects of vaccination and lockdown. In particular, the key element of Israel’s success has been suggested to lie in a high-dose vaccination rate that prevents the onset of a rebound in daily new cases on the rescission of the lockdown.

## 1. Introduction

At present, the world is still facing the crisis caused by the coronavirus disease 2019 (COVID-19) pandemic. The introduction of a lockdown cannot completely prevent the onset of a pandemic. Mass vaccination of the population is the most radical method of limiting its spread. Therefore, the Israel government authority started the 1^st^ and 2^nd^ vaccination operations against severe acute respiratory syndrome-coronavirus 2 (SARS-CoV-2) alpha variant on 19 December, 2020, earlier than most other countries. Because of the fairly small size of the Israeli population, cumulative doses exceeded 100% toward the end of March 2021, the fastest in the world. In parallel, the lockdown was also implemented. As a result, daily new cases drastically decreased from a peak of 10,000 to 500 at the end of March 2021. This successful experience should be shared in other countries. Hence, a prediction model that counts the number of infected people should be established given the data on vaccination dose rate and lockdown period. Few deterministic models based on the standard SIR model [1] have been reported [2] because it is usually applied at the initial stage of a pandemic when the number of susceptible individuals is close to the collective population. Conversely, a full simulation of the entire pandemic is needed to carefully consider the efficacy of the vaccine. To this end, the Agent-Based Model (ABM) has been recently investigated. The ABM [3],[4],[5],[6],[7] is a kind of stochastic approach and differs from mathematical models, such as SIR and SEIR, in that it can set up a virtual society that consisted a large number of autonomous decision-making and action actors (agent) with various attributes and can analyse complex social events involving interactions between actors. Based on the rules we set on a computer, agents spontaneously act while interacting in a given environment. Previous reports simulated the comparative and joint impact of COVID-19 vaccine efficacy and coverage together with non-pharmaceutical interventions (NPI) by taking the population of US cities as an example [6],[7]. They suggested that non-pharmaceutical interventions, such as masking and social distancing, among others, are still required to reduce infections against SARS-CoV-2 even after achieving a higher vaccine coverage and efficacy. In this method, a detailed setting is possible; however, many empirical constants are required because of its stochastic approach, and the calculation may be time-consuming depending on the scale. Moreover, a comparison was made based only on a cumulative basis. Prediction should be examined with factors such as in-patients and daily new cases, among others, by available observations.

Due to these difficulties in previous literature, a new deterministic approach, known as the Apparent Time Lag Model (ATLM) different from SIR, was developed [8]. The benefits of the model include the derivation of the final size equation and a simple analytic form of the fundamental reproductive number. In particular, ATLM should be emphasised based on the capability of calculating the trend of the effective reproductive number to foresee an infection turning point from growing to shrinking. These equations or correlations may provide a perspective on a pandemic of interest.

In the present study, ATLM was extended to be capable of evaluating the vaccine effects and NPI in Israel. The validity of the present model was shown using publicly available data. This model is characterised by a small number of empirical constants, which are all measurable.

## 2. Methods

### 2.1. Data

Infectious data of COVID-19 in Israel was referred to Worldometers [9] and vaccination statistics, and the reproductive number of COVID-19 was cited by Our World in Data [10]. The 1^st^ and 2^nd^ nationwide mass vaccination in Israel started on 19 December 2020, and the total dose rate per habitant exceeded 100% toward the end of March 2021. Lockdown was issued from 27 December 2020, to 5 February 2021 [11],[12]. Data for time lags *H* (period from infection until the polymerase chain reaction [PCR] reporting date), *V* (period of quarantine) and *T* (period from infection until quarantine) were set to 14 [13], 12 [14] and 14 days, respectively.

We adopted the SARS-CoV-2 infection data in Israel from 8 November 2020 to 31 March 2021 for analysis. Various data such as daily new cases, cumulative infections, currently isolated, currently infected and reproductive number were used to examine the numerical model. All vaccine doses during the 1^st^ and 2^nd^ campaigns are assumed to be Pfizer-BioNTech vaccine (BNT162b2 mRNA COVID-19 vaccine). Its real-world effectiveness was assumed 80% for the single-dose recipient at ≥10 days post-injection and similarly 95% for the double-dose recipient based on previous reports [15],[16],[17],[18],[19],[20].

### 2.2. ATLM with vaccine model

SARS-CoV-2 that caused the COVID-19 includes several delayed events in the spread of infection as follows:

1. Loss of infectious capability and extinguish within a month if no second infection occurs
2. The incubation time of about 5–7 days after being infected, in which the last 2 or 3 days are the infectious period.
3. Infected individuals are detected and recognised as PCR positives after approximately 2 weeks from infection.
4. PCR positives are mandatorily isolated and released after a certain time based on administrative guidance.
5. The effectiveness of vaccination would become fully effective in approximately 10 days after receipt of injection and have no waning nature.

Time delays considered in this study are listed in Table 1.

**Table 1.** Time delays adopted in ATLM

| Variable | Definition | Value | Reference | Remark |
| --- | --- | --- | --- | --- |
| T | Elapsed time until quarantine | 12–15 |  | T = L + F + G |
| L | Virus latent period | 5 | [21] |  |
| F | Elapsed time of pre-symptomatic state until PCR tested (temp. $\geq 37.5^\circ\text{C}$) | 5–7 | | |
| G | Go to the hospital for testing and test result reported | 2–3 |  |  |
| V | Isolation period | 12 | [14] |  |
| S | Elapsed time from infection until discharge/death | 26 |  | S = T + V |
| H | From infection until PCR testing results reported | 14 | [13] |  |

Therefore, the mathematics of the SARS-CoV-2 pandemic can be understood as demographics of progressive events with time delay. The Apparent Time Lag Model (ATLM) establishes governing equation consisting of a single discrete delay differential equation (DDE) in a normal manner. It differs from the conventional deterministic approach, such as the SIR model where the delay is implicitly included in the transmission rate with a reciprocal rate implying the first-order delay. ATLM provides a full simulation of a whole wave of the pandemic, whereas the SIR model expresses an entire solution as a combination of plural piecewise solutions to the respective small periods. Conversely, ATLM introduces a virtual collective population (*M*) instead of the real population (*N*) of a community in concern. As *M* is neither known a priori nor modelled so far, it must be estimated by the parameter fitting based on real-world infection data collected. Empirically, *M* is known to be much smaller than *N* [8], which matches the observation that the mobility of ordinary people in daily life is limited and the topological space is loosely closed. Thus, infections are likely to happen and cease in the area within the mobility. Random-based mass vaccinations are assumed to play a role in reducing *M*. Therefore, the following assumptions were considered in the analysis:

1. Every infected individual onset being PCR positive and eventually quarantined *T* days post-infection.
2. We count infections at large at the time *t* as *x(t)-x(t-T)* where *x(t)* is cumulative infected persons until t and infected individuals before *t-T* are all quarantined.
3. The portion of susceptibility may be expressed by *1-x(t)/M(t)*.
4. We assume the rate of new infection *dx(t)/dt* is proportional to the product of infections at large and the portion of susceptibility.
5. Individuals once infected are eventually removed and never infected again during the same pandemic.
6. There is no breakthrough infection.
7. We consider infection spread by the alpha variant only in the present study although observed data are a mixture of alpha variant and conventional strains with portions not negligible early in the time horizon.

Hence, the governing equation of ATLM becomes

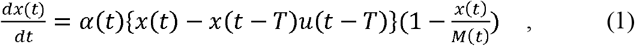

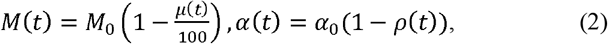

where *x(t)* indicates cumulative infected individuals; *dx(t)/dt*, rate of new infection; *α*_*0*_, transmission rate (1/day); *M*_*0*_, virtual collective population; *μ(t)*, effectiveness of vaccination (%); *ρ(t)*, dis-mobility of the people; and *u(t)*, step function. As the influence of vaccination corresponds to reduced susceptibility, we expressed the net collective population of a group as *M(t)* with consideration of the vaccination effectiveness. Then, the effects of lockdown were understood as the decrease in people’s mobility, resulting in decreased transmission rate *α*_*0*_ as in equation (2). Given the initial value of *x(0)*, Eqs. (1) and (2) were solved numerically using the 4^th^-order Runge Kutta Gill method with a time step of half-day and obtained *x(t)*.

Furthermore, other important infection-related parameters were calculated using *x(t)* as follows:

-Cumulative positive reported cases *Y(t)* appears in the delay of *H* days. Then,

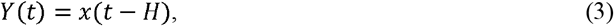

where *H* is the period from the onset of infection until the PCR test result report reaches the public health authority. *H* was assumed to be 14 days [13], i.e. days when the present data were collected. Equation (3) also indicates the number of cumulative isolated patients.

Similarly, daily new cases reported by the public health authority *dY(t)/dt* are daily new infections *dx(t)/dt* at H days before:

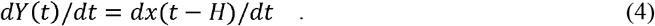

-Infections at large *W(t)* is assumed to be the sum of new infections in the period [*t–T, t*]:

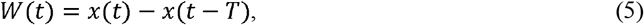

where *T* is a period during which an infected person remains infected at large (or until quarantined). It was assumed to be 14 days because *T = L + F + G*, where *L* is the incubation period of 5 days [21], *F* is the period of pre-symptomatic state before a temperature of 37.5°C becomes reached 5–7 days and G is the PCR testing and result reported 2–3 days, yielding 12(5 + 5 + 2) <*T*<15(5 + 7 + 3) with an average of *T = 14*.

-The number of isolated people *P(t)* consisting of hospitalisations, self-isolated at hotels and home and people who are still undecided on where to undergo treatment was counted based on the difference between those isolated and discharged using the following equation:

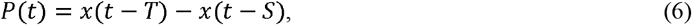

where *x(t-S)* is the number of recoveries or deaths. Denoting the isolation period as *V*, then *S* can be written as the sum of *T* and *V*, i.e. *S=T+V*. Israel’s Health Ministry decided to shorten the mandatory coronavirus isolation period from 14 to 12 days, provided that the patient had two negative tests from 12 November 2020 [14]. Then, *V=12* holds in Israel. Therefore, *S = T + V = 14 + 12 = 26*.

The effective reproductive number is understood as the average number of secondary cases to a primary case. ATLM defines the effective reproductive number *Rt(t)* as

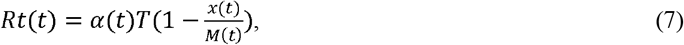

where *α(t)T* is the basic reproductive number [8].

In subsequent sections, predictions in Eqs. (3)–(7) will be compared with available data from Worldometer [9] and Our World in Data [10] and examine the present analysis method.

In summary, the values of delayed parameters adopted in the prediction are *H = T = 14, V = 12* and *S = 26*, which will be used throughout this study.

### 2.3. Vaccination modelling in Israel

Let the number of people vaccinated per 100 habitants be *A*, those vaccinated twice, *B*; and cumulative doses per 100 habitants *C*, then those vaccinated only once become *A–B* and *A + B = C* holds at any instance. When vaccination was completed, i.e. everyone has become a recipient twice, then *C* would be 200%.

Trends of data for *A, B* and *C* in Israel [10] are shown in Fig. 1. The target value of *B* is considered to be 5.5 million among 9 million citizens, which has been almost completed before the end of March. Let the vaccination efficacy for a single-dose recipient be β and that of double-dose one be γ, then the average efficacy *μ(t)* of a group consisting of both *A* and *B* is written as follows for a single-type vaccine:

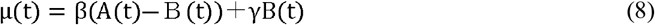

**Fig. 1.**
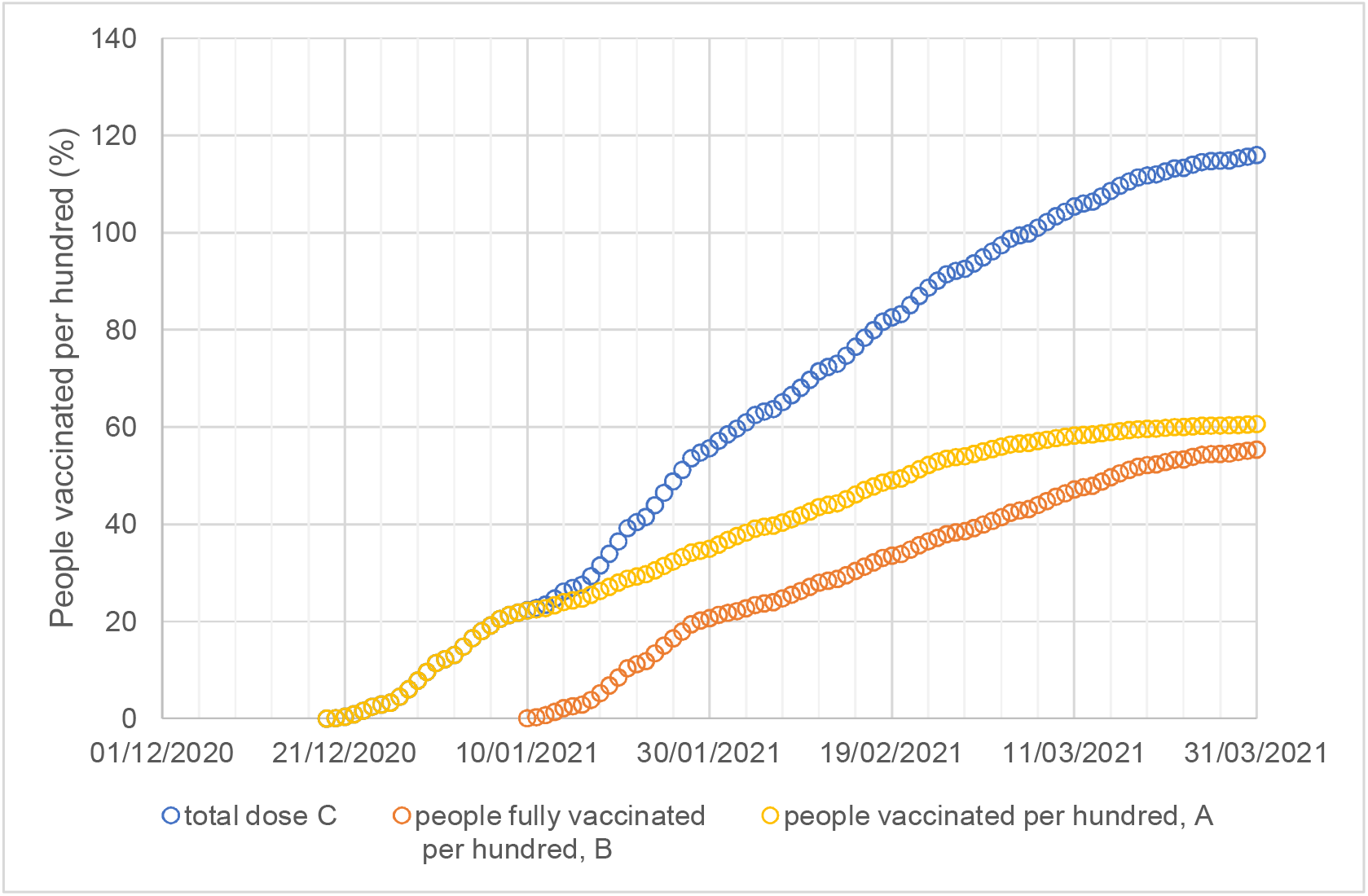
The number of people in *A* and *B* per 100 population and the total vaccine dose per 100 habitants *C* in Israel. The abscissa represents the date, 25 October 2020, when the calculation started. *A, B* and *C* are observed data in which *A* and *B* represent the number of people vaccinated per 100 population and those vaccinated twice in Israel. *C* is the total dose per 100 habitants. The axis shows the number of people or doses per 100 population. The vaccination started on 19 December 2020 onward. The total doses exceeded 110 at the end of March 2021.

The type of virus in the 3^rd^ wave of the `pandemic` in Israel is mainly the alpha strain [15]. The efficacy of the BNT162b2 mRNA COVID-19 vaccine (Pfizer-BioNTech COVID-19 vaccine) in a nationwide mass vaccination campaign in Israel is reported in peer-reviewed articles [18],[19],[20]. Based on these, we adopted β = 0.8 and γ = 0.95 in Eq. (8) and assumed the vaccine was activated after 10 days for both single and double injection recipients.

The data plot of *μ(t)* (Eq. (8)) in blue circles and its polynomial approximation Eq. (9) in broken red line are presented in Fig. 2.

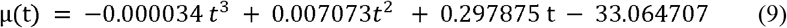

**Fig. 2.**
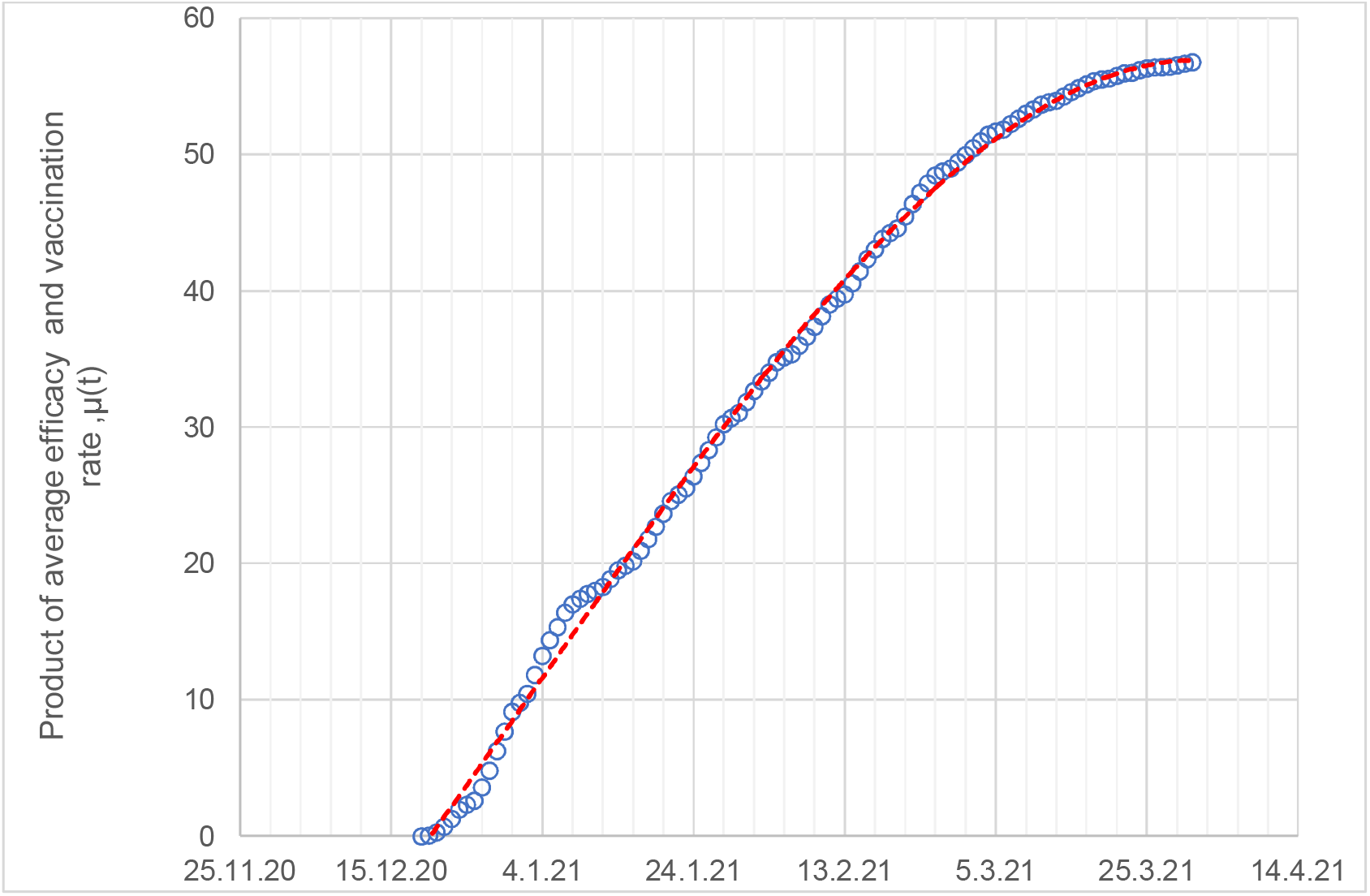
Presumed collective vaccine effectiveness (in case of no delay) inferred from the observed data by Eq. (8) for a collective population composed of recipients mixed up with single- and double-dose vaccinations. The axis represents the product of average vaccine efficacy multiplied by the vaccination rate (referred to as collective vaccine effectiveness [CVE]). The vaccination started on 19 December 2020.

Since the vaccine effectiveness becomes activated in *ξ* days post-injection, *μ(t)* in Eq. (9) was replaced by *μ(t-ξ)*. Based on previous literature [16] [17], we assumed *ξ = 10* in the computation.

It is observed that the vaccination speed was quick and the CVE reached approximately 40% 60 days after starting the vaccination.

### 2.4. Lockdown modelling

Lockdown suppresses human contact to reduce infections in society. The Israeli government executed a national lockdown as a necessary step to block the increased morbidity [10] on 27 December 2020 (63days after 25 October) for 2 weeks with an option of extending it. Actually, a national lockdown was extended until 5 February 2021 (103days) due to delayed turnaround time from the health and economic crises on 31 January 2021 (98 days) [12].

We modelled the lockdown effects by a sudden decrease in the transmission rate as shown in Fig. 3. *(1-ρ(t))*_*min*_ was set to 0.72 as a result of parameter fitting. Other parameter values determined after fitting were *x(0)* = 650, *α*_*0*_ = 0.117 and *M*_*0*_ = 1,500,000, respectively. First, their values were identified as the 1^st^ approximation using data in an early phase of the 2^nd^ wave (for details, see Appendix-A). Further, these values were graphically tuned so that they should be consistent with those of other measurements, such as the number of infections at large, in-patients and the trend of the reproductive number. Table 2 summarises calculation conditions and actual event sequences.

**Table 2.** Calculation conditions and actual event sequence

| Item | Variable | Unit | Value | Remark |
| --- | --- | --- | --- | --- |
| Transmission rate | $\alpha_0$ | 1/day | 0.117 | |
| Initial susceptible | $M_0$ | - | 1,500,000 | |
| Initial infected people | $x(0)$ | - | 650 | 25 October 2020 |
| Delay until quarantine | $T$ | day | 14 | |
| Period of quarantine | $V$ | day | 12 | |
| Delay until discharge | $S$ | day | 26 | $S = T + V$ |
| Delay until reporting<br>PCR positive result | $H$ | day | 14 | |
| Start of calculation | $t$ | day | 0 | 25 October 2020 |
| Start of investigation | $t$ | day | 14 | 8 November 2020 |
| Start of vaccination | $t$ | day | 55 | 19 December 2020 |
| Start of lockdown | $t$ | day | 63 | 27 December 2020 |
| End of lockdown | $t$ | day | 103 | 5 February 2021 |
| End of calculation | $t$ | day | 157 | 31 March 2021 |

**Fig. 3.**
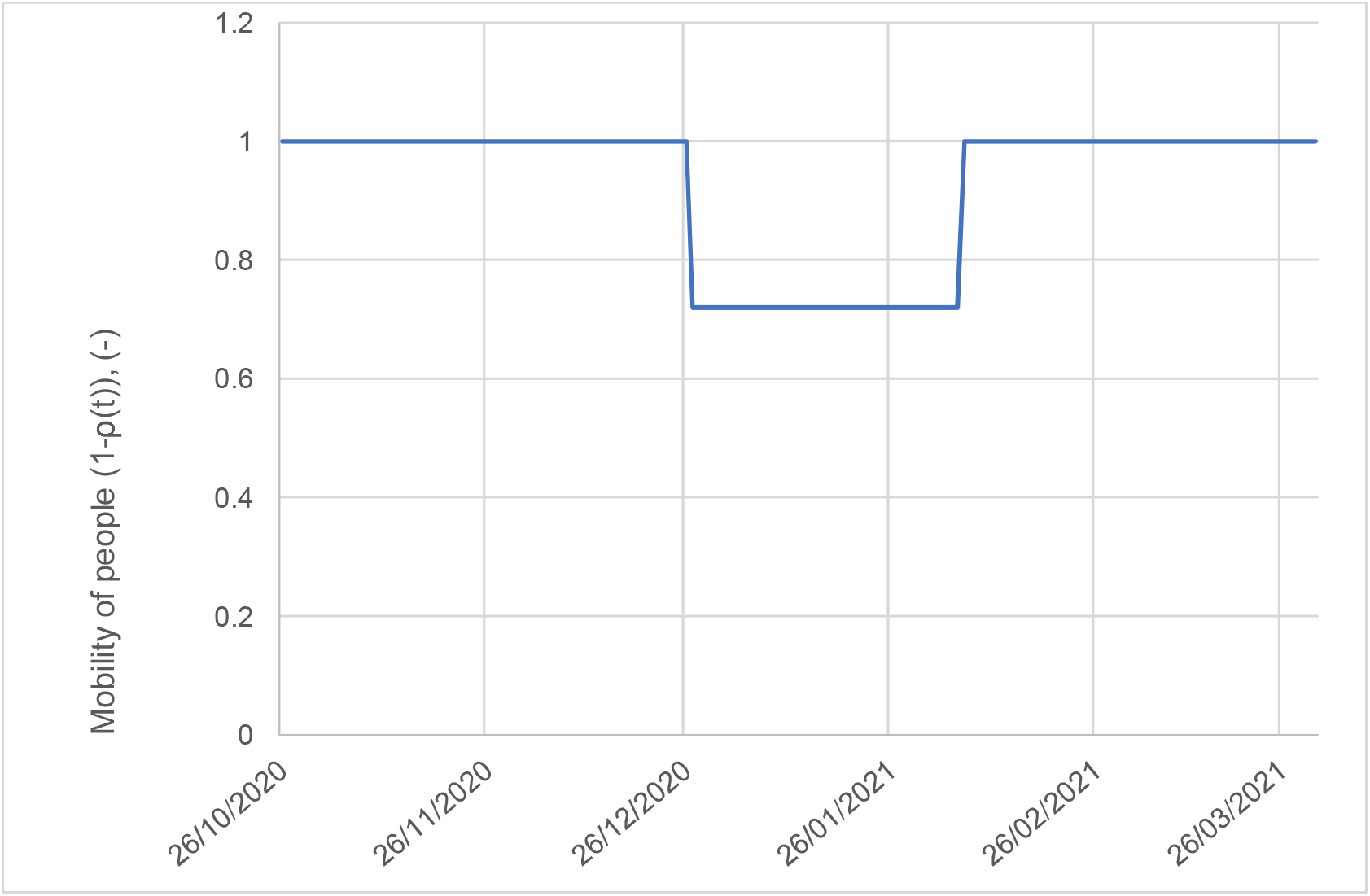
Lockdown modelling. The axis represents *1-ρ(t)* in Eq. (2). Lockdown was operated from 27 December 2020, to 5 February 2021. Its effect is anticipated to become apparent on 10 January 2021, and 19 February 2021, because of a 14-day delay in reporting.

## 3. Results

### 3.1. Daily new cases and total cases

Daily new cases were obtained from the Worldometer data source [9] first for comparison with the prediction by ATLM equipped with vaccination and lockdown models. The study was conducted from 8 November 2020, to 31 March 2021, covering a whole span of the third wave of the COVID-19 `pandemic` in Israel in which mass vaccination and a nationwide lockdown were involved. We regarded the term ‘daily new cases’ in Worldometer as daily new positives observed behind a new infection based on *H* days (14 days). The computation started ahead of 8 November 2020, by *H* days, i.e. 25 October 2020, the original calculation time.

Figure 4 compares the prediction using the observed data. Both are expressed on a 7-day average basis to prevent fluctuation using the difference in the number of PCR tests in a week. The prediction is a 7-day average of *dY/dt* (Eq. (4)). The following important findings were observed:

**Fig. 4.**
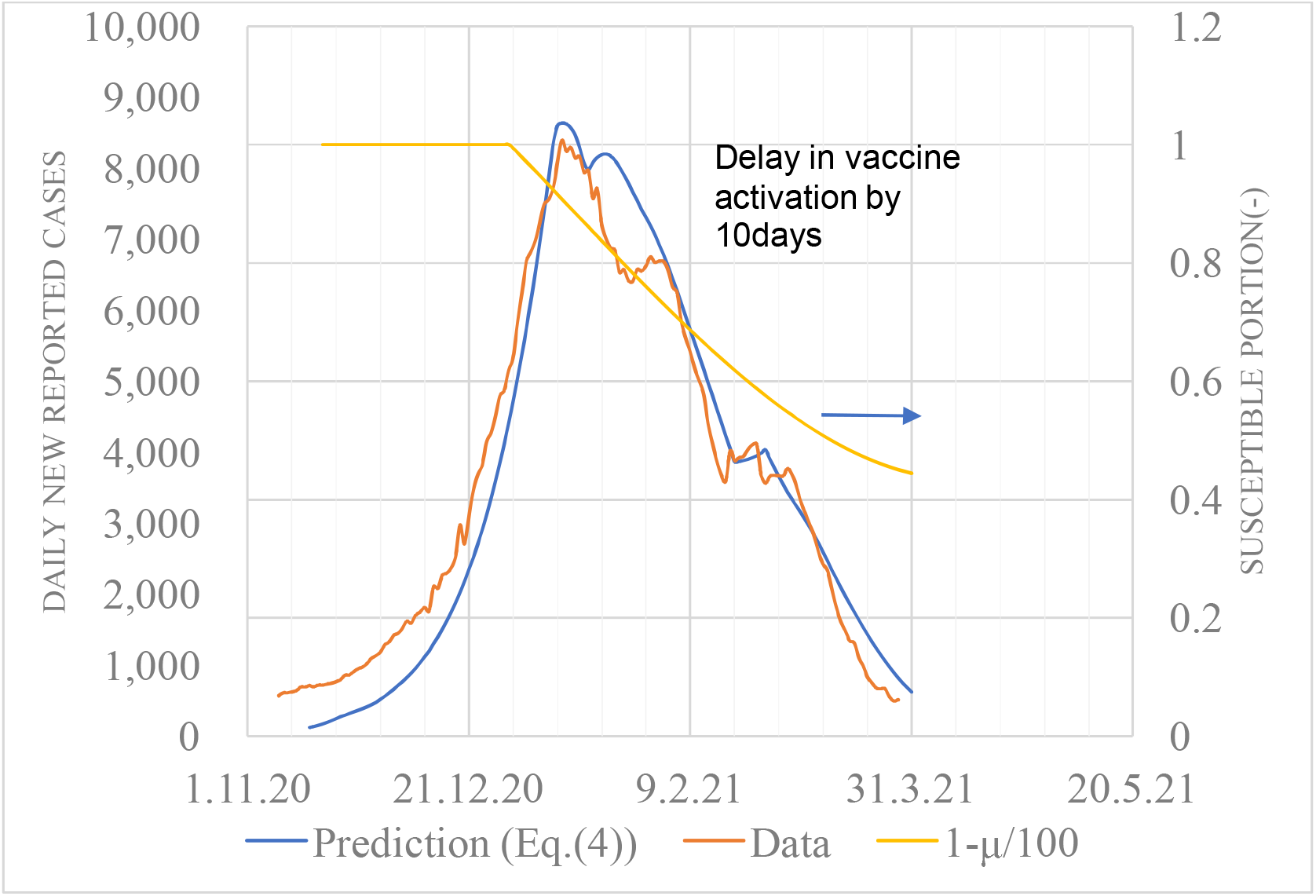
Daily new cases based on the observation and prediction. Red line: data, blue: prediction, yellow: reduction ratio of susceptible due to vaccination *1-*μ*(t)/100* referring to the right axis. Left axis: the number of newly infected people, Abscissa: the origin of computational time is 25 October 2020, 14 days ahead of 8 November, the start of the 3^rd^ wave of the pandemic in this study. Both data are displayed as a 7-day average of backward movement. The 3^rd^ wave of the COVID-19 pandemic in Israel was regarded to start from 8 November 2020, to the end of 31 March 2021. A nationwide mass vaccination started on 19 December 2020. Its effectiveness was revealed behind 10 days. A lockdown was implemented between 27 December 2020, and 5 February 2021. The calculation shows approximately 7 days *dY(t)/dt*, which was carried out using Eq. (4).

1. Impacts at the start and the release of lockdown are apparently observed in data on both dates as 10 January 2021, and 19 February 2021, respectively, whereas the actual dates when the public health interventions were 27 December 2020, and 5 February 2021, respectively [12]. The former reduces the infection rate resulting in the appearance of a peak, whereas the latter produces a plateau as a result of an antagonist between the rebound of the lockdown rescission and the vaccination efficacy. These characteristics are found to have been well simulated. This fact validates the assumption that *H* = 14 (infection becomes apparent in a 14-day delay). Both observed data were simulated within 10% error in terms of both time and magnitude.
2. At the early stage of the pandemic, the prediction is found to underestimate the observed data. This fact can be interpreted as follows. The observed data contain tailing effects of the preceding 2^nd^ wave, which were not considered in the present study.
3. The above-mentioned plateau was not reproduced during computation. This might be caused by an initial additional lockdown intervention on 8 January 2021, as disclosed in the recent literature [22]. Overall, however, the simulation of the entire trend of daily new cases was satisfactory.

Figure 5 presents cumulative positives *Y(t)* (Eq. (3)). The overall agreement was achieved between the prediction and observed data. Underestimation was obtained from the fact that the observed data contain the tailing effect of the preceding 2^nd^ wave, which was not considered in the present study. Remarkably, the number of cumulative positives attained was approximately 500,000, which is approximately 1/3 of the virtual collective population *M*_*0*_. Then, 2/3 of the total population remains uninfected because the term {·} of RHS in Eq.(1) governs infection phenomenon toward the endemic rather than the term 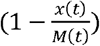 due to the lower transmission rate. Our previous study [8] showed this may become dominant provided that *α*_*0*_*T* < 2.5, whereas the value in the present study is 1.64.

**Fig. 5.**
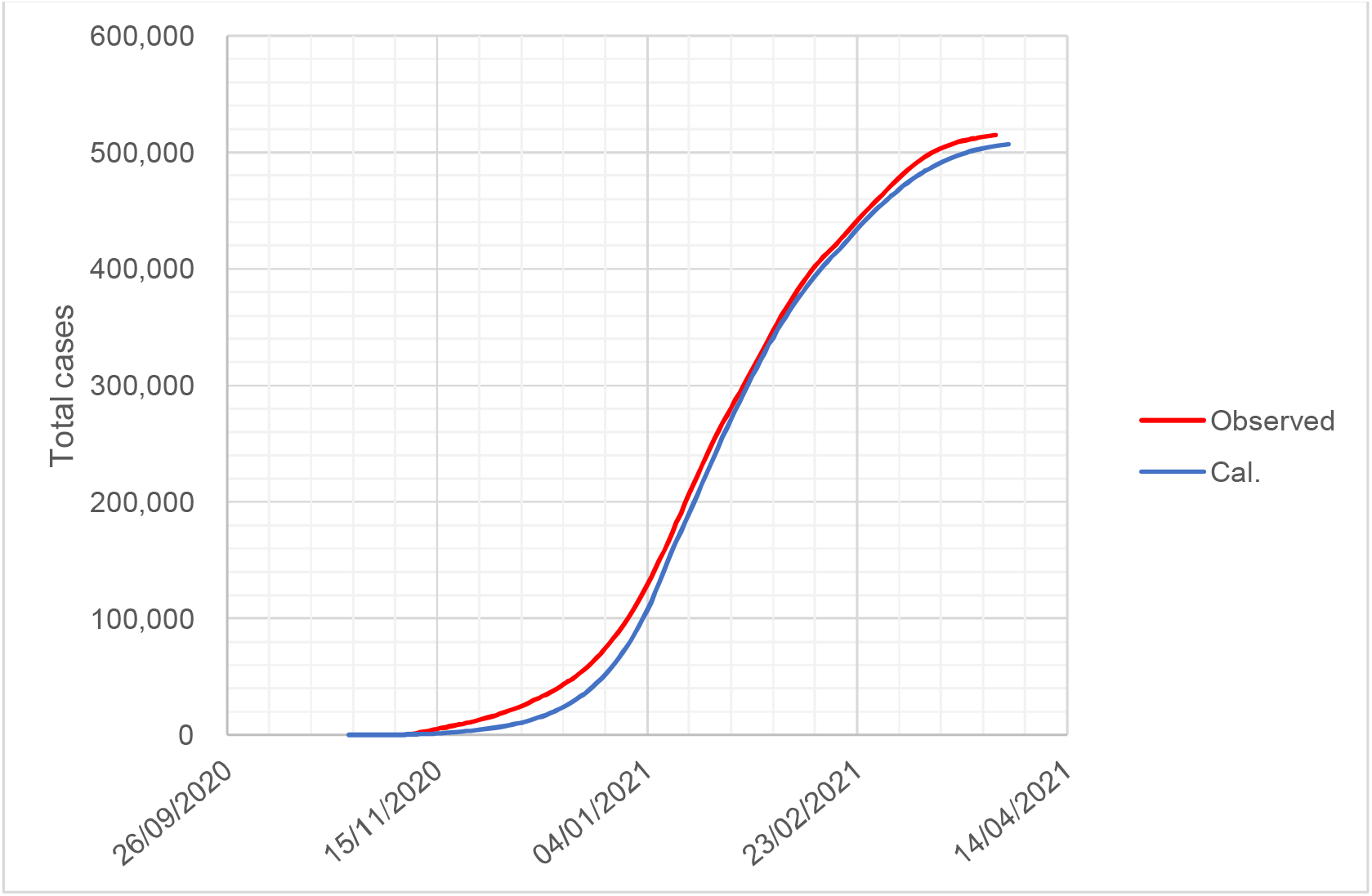
Trends of cumulative positives. Equation (3) was used to calculate cumulative positives, *Y(t)*

### 3.2. The number of isolated patients

The number trend of isolated patients is presented in Fig. 6. Although the calculation using Eq. (6) overestimates the actual data by 15% at a peak on 20 January 2021, the overall trend is simulated fairly well and also proves the assumption that *T = 14* in the model that is correct because of a larger value of *T*, resulting in overestimation and vice versa for a smaller one.

**Fig. 6.**
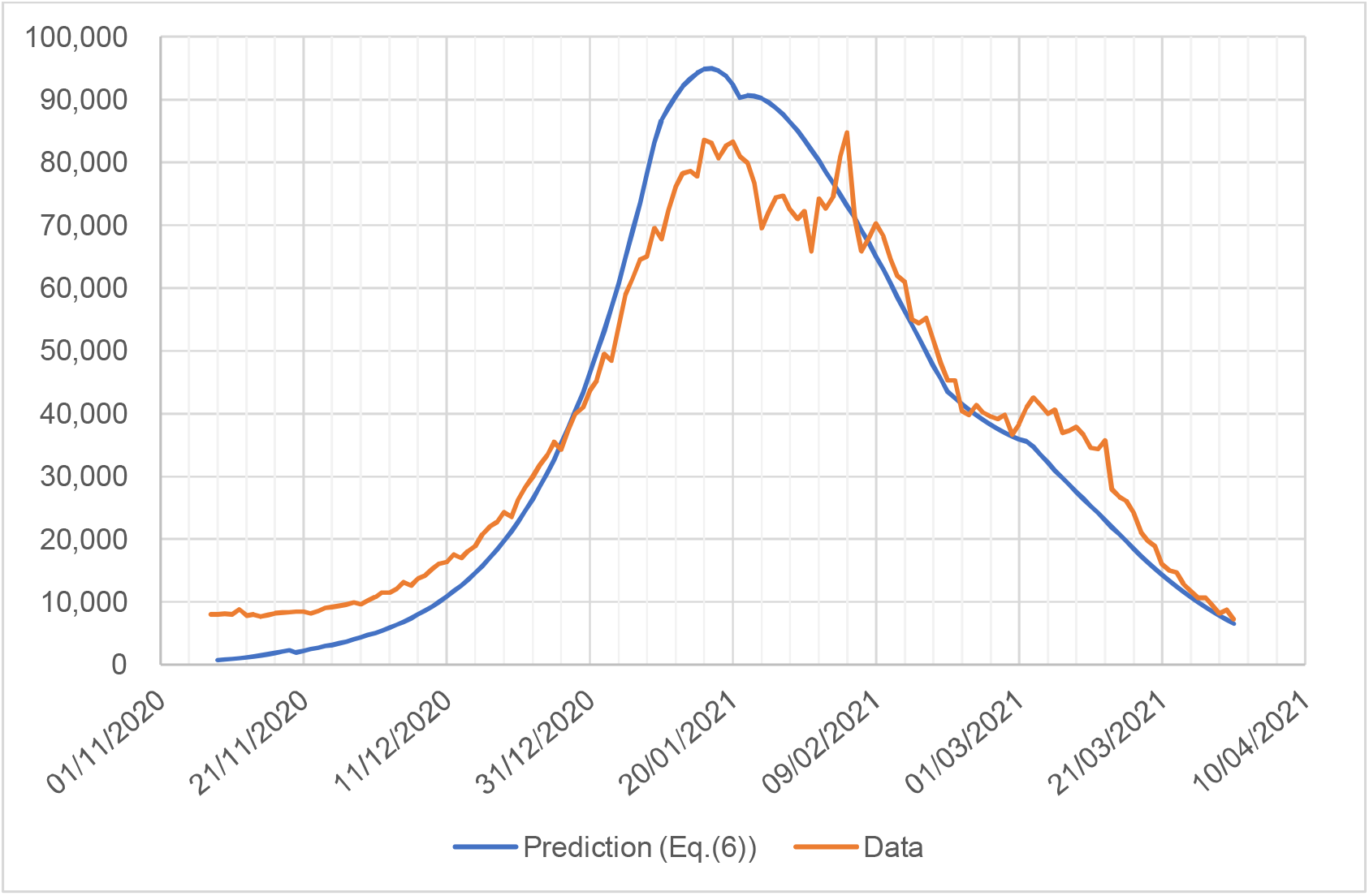
Trend of currently isolated patients. The red line indicates data cited from Worldometer with a notation of ‘currently infected’ [9]. The blue line is the prediction using ATLM. Coincidence in timing implies the assumption *T = 14* is proper. The time to provide the peak is behind that of daily new cases (Fig. 4) based on a week.

### 3.3. Reproductive number

Figure 7 compares the prediction using Eq. (7) with data [10] based on the reproductive number (Rt(t)). The *Rt(t)* data in Our World in Data have been compiled using the method described in the literature [23] and contain real-time estimates of the effective reproductive number. Their *Rt(t)* estimates for COVID-19 in 124 countries worldwide are provided and used to assess the effectiveness of non-pharmaceutical interventions in a sample of 14 European countries. Thus, the data could be most reliable at this moment.

**Fig. 7.**
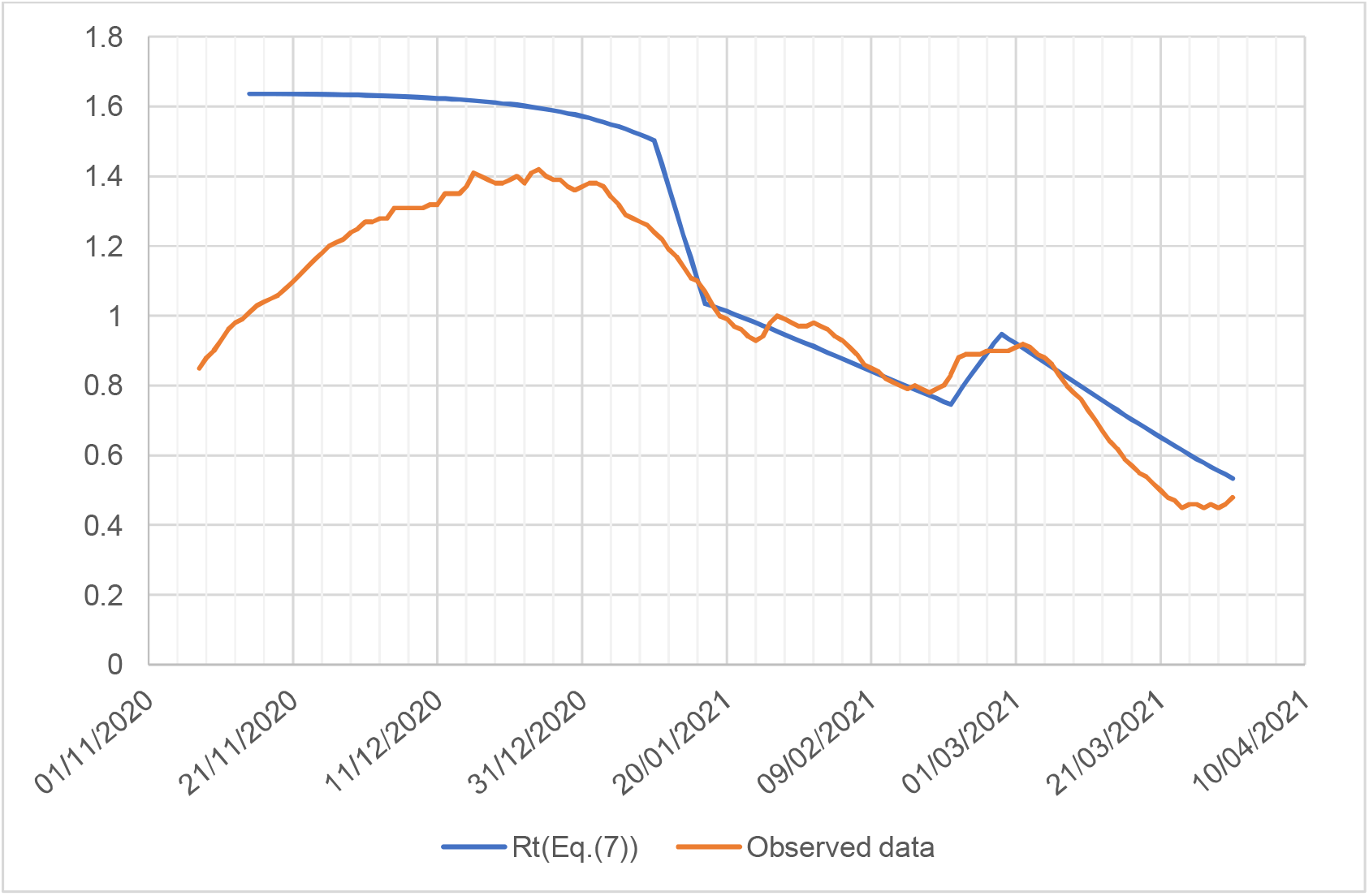
Effective reproductive number. Observed data were cited from our world in data [10] and Eq. (7) was used for calculation.

Aside from an early phase, prediction for the 3^rd^-wave pandemic in Israel coincides well with estimates from the observed data, especially during the vaccination and lockdown periods. The discrepancy between data and calculation in the early phase could be accounted for based on the co-existence of the aforementioned tail of the 2^nd^-wave pandemic.

In the early phase, the reproductive number is a compound of the 2^nd^ and 3^rd^ waves, which may be estimated by a weighted average *Rt*_*mixed*_ = *Rt*_*2nd*_*(0.83) + *Rt*_*3rd*_* (0.17) on 15 November. In Fig. 7, substituting the tail part value of *Rt*_*3rd*_ for *Rt*_*2nd*_ would provide *Rt*_*2nd*_ = 0.5. Then, with *Rt*_*3rd*_ = 1.64, *Rt*_*mixed*_ = 0.5*(0.83) + 1.64*(0.17) = 0.70. As the tail shrinks, *Rt* grows, which could explain such a tendency of the observed data in the early phase.

The basic reproductive number *α*_*0*_*T* for Israel was estimated using the ATLM as 0.117 × 14 = 1.64, which is smaller than those evaluated by [24] in the USA and eight European countries using data without pharmaceutical intervention. The reason for its difference remains unknown yet; however, the accumulation of data with pharmaceutical intervention would be recommended for future work.

### 3.4. The number of infections at large

Infections at large should be recognised as an important variable to determine when the pandemic will end, a finding much more accurate than daily new cases because the time to reach *W(t) = 0* rigorously indicates the end of a pandemic.

Next, infections at large may be obtained using data processing as follows. The substitution of daily new cases *Y(t)* to RHS of Eq. (5) would yield *W(t-T). W(t)* would be obtained by shifting the *W(t-T)* profile leftward by *T* along the time axis. To confirm the validity of the obtained *W(t)* in this method, an agreement was confirmed with a numerical prediction with the precision that has already been examined through arguments described in the previous section. As shown in Fig. 8, an excellent agreement is observed between the two, suggesting the correctness of the data processing mentioned above. The *W(t)* peak to *dY(t)/dt* ratio was evaluated to be approximately 13 times.

**Fig. 8.**
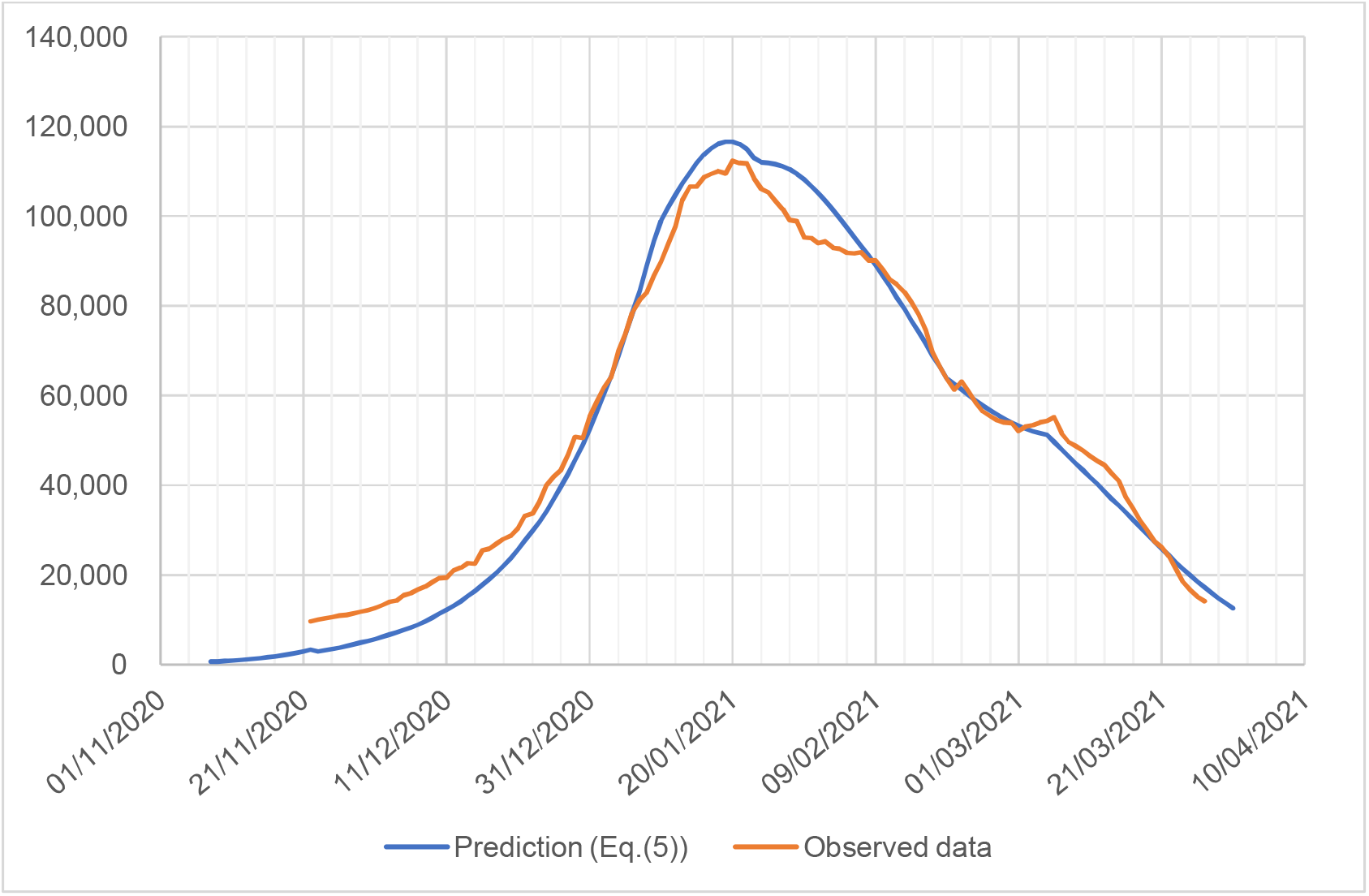
Infections at large. The red line is processed from data, whereas the blue line is from prediction.

### 3.5. Estimation of the separate effects of vaccination and lockdown

Based on the confirmed epidemic model, parameter survey calculations were conducted for the following four scenarios:

1. Without vaccination, without lockdown: case C *μ* = *ρ* = 0
2. With vaccination, without lockdown: case D *μ* ≠ 0, *ρ* = 0
3. Without vaccination, with lockdown: case B *μ* = 0, *ρ* ≠ 0
4. With vaccination, with lockdown: case A (real intervention) M ≠ 0, *ρ* ≠ 0

Results are presented in Fig. 9.

**Fig. 9.**
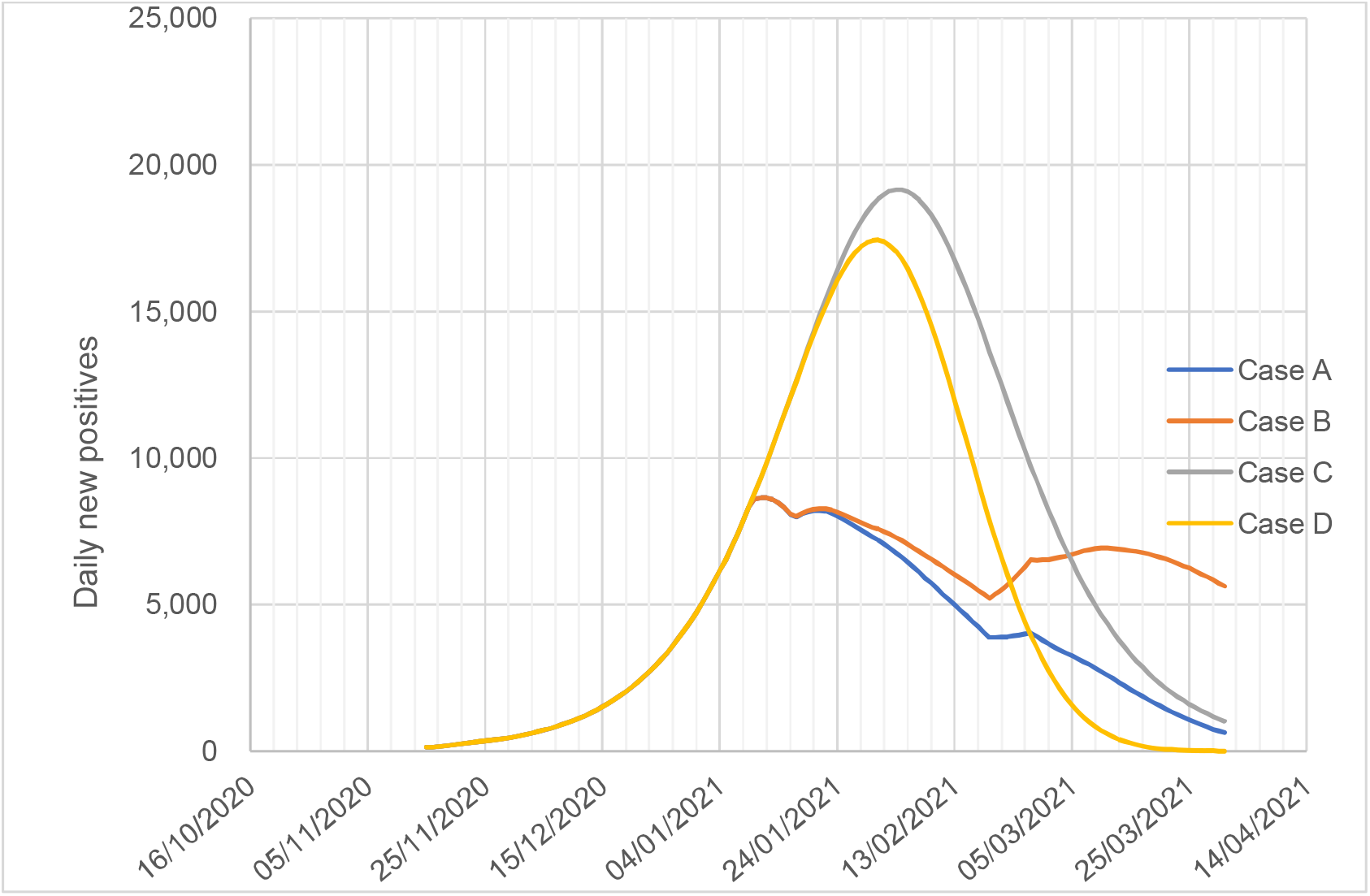
Predicted daily new cases for various combinations of public health interventions with or without vaccination and lockdown. Axis: daily new cases, Abscissa: time (date), case A: With vaccination, with lockdown, case B: Without vaccination, with lockdown, case C: Without vaccination, without lockdown, case D: With vaccination, without lockdown.

Without lockdown, the blue line (A) would have shifted to the yellow line (D). If the vaccination dose rate had been slower, then, the blue line (A) would have approached the red line (B). In general, the lockdown has decreased the transmission rate, whereas vaccination accelerates the convergence of a pandemic.

Next, the separate effect of either vaccination or lockdown was investigated.

We will introduce *D/C* and *B/C* ratios in which *B, C* and *D* mean daily new cases of case B, C and D. Without lockdown, the peak value would increase 2–2.5 times larger than in the real one denoting *C/B*. Without vaccination, the `pandemic` would decelerate in a later stage *C/D* and even worse on the earlier release of the lockdown.

Figure 10 presents the joint impact of both ratios.

**Fig. 10.**
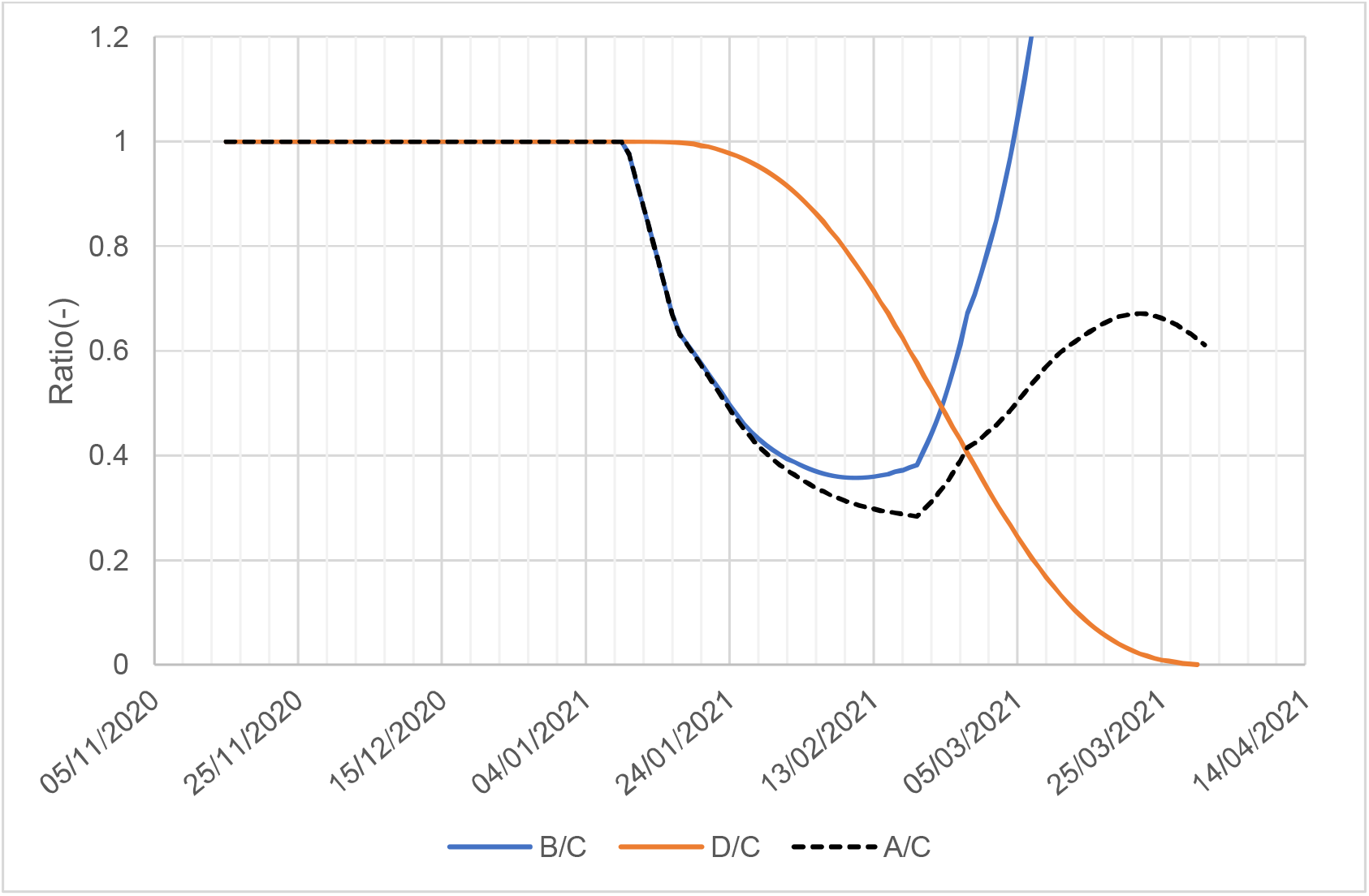
Separate effects of vaccination and lockdown reduced the infection. The *B/C* ratio (blue line) denotes the lockdown effects without vaccination, the *D/C* ratio (red line) represents the vaccination effect without lockdown and the *A/C* ratio (black broken line) shows a real combined effect.

Here, the ratio means a relative infection rate to *C*. The smaller the ratio, the lower the infection rate. As vaccinations proceed, the *D/C* ratio monotonously decreases. Conversely, the *B/C* ratio of the lockdown has a minimum due to rebound during the rescission of lockdown. It is observed until 22 February 2021, the impact of lockdown exceeds that of the vaccination. Thereafter, the effects of vaccination are superior to lockdown. The combined effects of *A/C* are presented in a broken line, indicating that rebound suppression may be to some extent caused by vaccination. A period (25 January 2021–25 February 2021) of synergy was observed when the black broken line has the smallest ratio among the three. It is inferred that the faster the dose rate, the smaller the rebound magnitude, which would contribute to the earlier ending of the pandemic.

### 3.6. Sensitivity analysis

Time delay constant *T* may vary from country to country depending on many factors, such as the type of virus strain, public health regulation, intervention, governmental economic policy, medical resources and knowledge of the virus strain and variant, among others. Sensitivity analysis was conducted to determine what *T* value is appropriate for the present Israeli COVID-19 pandemic. Taking 16, 15, 14, 12 and 10 days as candidates, the transmission rate *α*_*0*_ and initial virtual collective population *M*_*0*_ were estimated using the method in Appendix-A with data corresponding to the growing phase of the pandemic before lockdown or starting the mass vaccination. Numerical simulations using Eqs. (1) and (2) were conducted from the beginning until the end of the pandemic and compared with the data. This calculation should be started on a different day based on *T*. The number of initial infectors *x*(0) in each case was defined as a value on *T* days before 8 November so that the new reported 7-day average delay (=238) should be common to all on 15 November 2020.

Results are presented in Table 3 and Fig. 11.

**Table 3.** Dependency of the time delay *T* on estimated values of *α*_*0*_, *M*_*0*_ and *x(0)*

| VARIABLE | UNIT | VALUE |  |  |  |  |
| --- | --- | --- | --- | --- | --- | --- |
| $T$ | (DAY) | **16** | **15** | 14 | 12 | 10 |
| $\alpha_0$ | (1/DAY) | **0.0995** | **0.104** | 0.108 | 0.119 | 0.135 |
| $M_0$ | (-) | **2,000,000** | **1,850,000** | 1,670,000 | 1,440,000 | 1,350,000 |
| $x(0)$ | (-) | **1,705** | **1,600** | 1,520 | 1,315 | 1,080 |
| $\left(\frac{dy}{dt}\right)_{t=20.5}$ | (1/DAY) | **251** | **251** | **251** | **251** | **251** |
| $\alpha_0 T$ | (-) | **1.592** | **1.560** | 1.512 | 1.428 | **1.35** |
| $\left(\frac{dY}{dt}\right)_{\text{PEAK}}$ | (%) | **104** | **102** | **100** | **100** | **98** |

**Fig.11.**
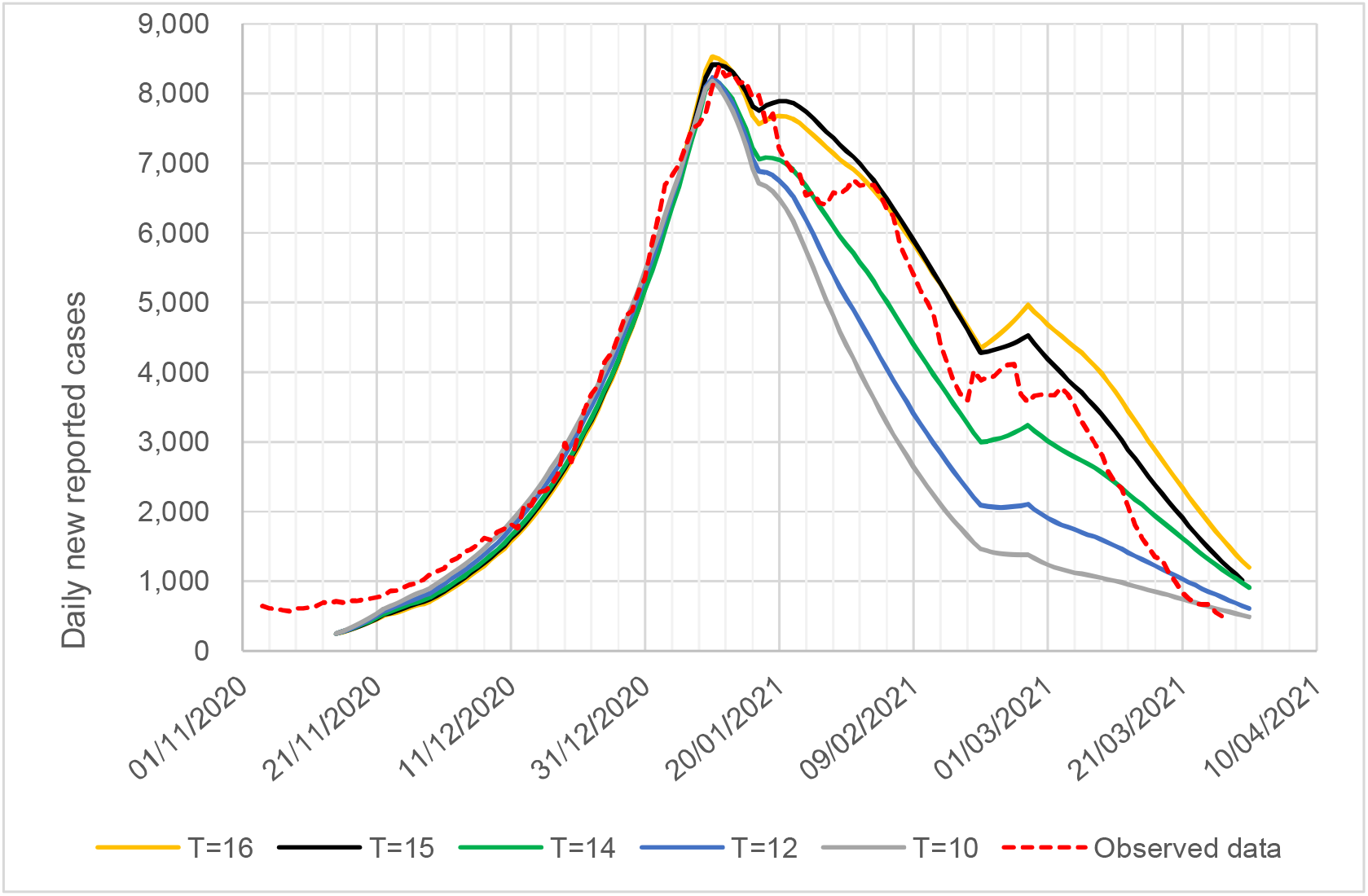
Sensitivity analysis of the time delay *T*

It is observed as *T* decreases and *M*_*0*_ decreases, whereas *α*_*0*_ and *α*_*0*_ *T* increase. In Fig. 11, each calculation shows an excellent agreement with the data trend until the peak, except at the very early stage, but is underestimated in most of the descending stages after the peak, except for T = 16. One possible reason for the underestimation may be obtained from the fact that senior citizens received injections first before the younger generation whose social activity is much higher. However, this model is not sophisticated as to accommodate such an inhomogeneity. As a result, the predicted effectiveness of the society may overestimate the real one and hence underestimate infections at an early phase of vaccination. Among the four cases, the discrepancy was found smallest in the case *T* = 14. Noting this fact and empirical knowledge on the allowable range of time delay (Table 1), T = 14 was adopted. Furthermore, these are finely tuned so that they should be consistent with the other measurements, such as the number of infections at large (Fig. 8), in-patients (Fig. 6), and trend of effective reproductive number (Fig. 7).

The final results are presented in Table 2. Moreover, the portion of daily new reported cases during the 2^nd^ wave of the pandemic on 15 November was estimated to be 1/3 of the total population (observed data).

## 4. Discussion

### 4.1. Primary findings

The ATLM equipped with a newly developed vaccination and lockdown model simulated a whole trend of various infection variables for the third wave of the COVID-19 pandemic in Israel within a 15% error. In particular, the number of isolated PCR positives and infections at large was separately evaluated with high resolution. Moreover, in the mid-span and after the pandemic, the prediction of an effective reproductive number using Eq. (7) derived from ATLM agreed in the period of the later half of the pandemic with real-time estimates in Israel [10] using this method [23]. It provides estimates of *Rt* for COVID-19 in 124 countries worldwide, which are used to assess the effectiveness of non-pharmaceutical interventions in a sample of 14 European countries. A huge difference appearing during the initial phase of the pandemic was quantitatively explained, i.e. it was affected by the tailing part of the preceding pandemic that occupies 2/3 of the total population.

These facts confirm the present vaccination model whose concept is to reduce the virtual collective population based on the acquired vaccination effectiveness. Conversely, most previous studies mainly using ABM disclosed to date were limited to the total and daily new cases [4],[5],[6] and were unlikely to show prediction of the trend of the effective reproductive number.

A separate effect of vaccination and lockdown was attempted to be explored. Although lockdown most effectively suppresses the infection rate in the ascending phase of the pandemic, inducing a rebound on its release is inevitable. Vaccination is found to be a strong tool to suppress it. To make this effective, it was inferred from parameter survey calculations that the vaccination dose rate should be sufficiently rapid to achieve at least 40% effectiveness during the rescission of a lockdown. This effective value was found consistent with the herd immunity value drawn from a reproductive number analysis.

Additionally, the data processing method to extract infections at large from the total cases was clarified. These infections at large *W(t)* are a decisive index to estimate the end of the pandemic when *W(t)* becomes zero, occurring behind the time when daily new cases become zero and yielding a conservative measure to plan public health prevention and control the pandemic.

A previous study [8] reported that the herd immunity level is evaluated as 1 − 1/ *αT* =1 − 1/(0.117 × 14) = 0.39. Figure 2 indicates the herd immunity level is established on 13 February 2021. Therefore, a collective vaccine effectiveness of 0.4 is found to be adequate in Israel to suppress infections.

The transmission rate *α*_*0*_ obtained should be discussed. The Israeli 3^rd^-wave COVID-19 pandemic is compared with that in Tokyo, with a similar population size of 14 million to Israel’s 9.8 million. The prevalent strain in the 4^th^-wave pandemic in Tokyo was an alpha strain, a finding similar to the 3^rd^-wave pandemic in Israel.

*α*_*0*_ was evaluated to be 0.09 in Tokyo using the same approach, i.e. ATLM with *T* = 14 [25] but 0.117 in Israel. It may come from the difference in pandemics with and without vaccination. Anyway, the difference between the two is not large, and thus, the currently obtained *α*_*0*_ is reasonable. For more information, the conventional strain during the 1^st^-wave of the pandemic and the delta strain in the 5^th^-wave pandemic both observed in Tokyo have 0.164 [8] and 0.121 [25], respectively.

In the present study, ATLM did not include the asymptomatic patient. Nonetheless, the prediction succeeded in explaining the profile of various observed data trends within a tolerable range of errors, especially for the number of isolated individuals and infections at large. This fact suggests that most symptomatic patients if any might have been non-infectious in Israel.

### 4.2. Limitations and future work

Limitations of the present method are as follows: First, the present vaccination model presumed the random order of receiving vaccines based on age and sex in a community in concern; however, senior citizens received vaccines on a priority basis and then middle-aged and younger generations follow in Israel. This might have increased the positive number more than the present numerical results because the active generation is likely infected due to their higher human contact.

Second, we assumed no waning of vaccination immunity. This assumption is proper if the vaccine injection rate is sufficiently rapid to induce a waning of immunity.

No available data have been established on the waning characteristics for the alpha variant currently under consideration. However, noting that herd immunity seems to have been established in 3 months, the above assumption is proper in the present calculation. However, this assumption cannot be applied to the omicron strain because the waning is faster than ever. Therefore, our future work should develop a method to evaluate the effectiveness in a community with the waning of immunity after considering the vaccination.

Third, the present ATLM is a homogeneous model for average individuals and disregards their differences among generations. The associated parameters are the transmission rate, virtual collective population, characteristics of vaccine-waning immunity and order of vaccine injection by age. Considering the difference between generations, a heterogeneous model is needed, which is composed of generation-wise equations with relevant parameter values, which will be again our future task.

The present vaccination model is established based on the efficacy data of the BNT162b2 mRNA COVID-19 vaccine (Pfizer-BioNTech COVID-19 vaccine). It should be extended to accommodate other COVID-19 virus strains, variants and vaccines.

Fourth, generally, deterministic approaches including the ATLM assume a homogeneous model to calculate the spread of infection. Namely susceptible as well as infectious people are supposed to be well mixed in a society at any time. In the real world, however, infection begins spreading at a specific time and place in the society and majority people move along their routine courses in daily life. Then, in a community with small size population, the routine courses may reach all through the community and the infection can spread throughout the community. To the contrary, in a community with large size population and/or area, the infection spread will not reach throughout and tends to be localized because infectious period of the virus is limited. Such population heterogeneity would limit the magnitude of infection spread. In another words, ordinary persons move around in a limited portion of the community and the possibility of encounter with those who live far away is quite small. Then, the effective size of population in a community *M* in terms of infection spread may be much smaller than real population size. Unfortunately, nobody has not found any model yet to evaluate the size of *M*. Therefore, we estimated *M* empirically based on the observed infection data in a growing phase of the pandemic of interest.

In summary, this is the first study of a full simulation during the 3rd wave of the COVID-19 pandemic in Israel accompanied by a mass vaccination campaign and simultaneous lockdown. Existing data and policymaking elsewhere would be aided by including the present model. Potential users of the present model are advised to reassess the values of delayed constants *T, H, V* and *ξ* in the model when the application to other countries is attempted because they are influenced by the difference in medical regulations or practices.

## 5. Conclusion

A simple numerical epidemic model ATLM was extended to simulate an entire trend of various infection-related variables by considering the vaccination campaign and simultaneous lockdown. The model is described by a single discrete DDE with explicit inclusion of the time lag of infection isolation. It provides a full simulation of various infection variables in an entire period from the initial infection through the end-point with a small number of fitting parameters. The time series data of both vaccine dose ratio and lockdown period were used in this model. The model was examined within 15% of errors compared with Israel’s various COVID-19 pandemic trend data such as daily new cases, the number of isolated people and infections at large and effective reproductive number. Moreover, parameter survey calculations for several scenarios have clarified that a synergy effect exists between vaccination and lockdown. In particular, Israel’s success in the avoidance of the onset of the rebound lies in a high-dose rate in mass vaccination campaign so that herd immunity can be achieved before rescission of the lockdown.

## Data Availability

We used worldometer and our world in data for COVID-19 infection data in Israel as source data both of which are publicly accessible.

https://www.worldometers.info/coronavirus/country/israel/

https://ourworldindata.org/covid-vaccinations

## Data availability

We used time-series data of COVID-19 from 8 November 2020, to 31 March 2021, in Israel [9],[10].

## Author contributions

M. Utamura developed and verified the epidemiological model. S. Kirikami developed the parameter identification method. M. Koizumi examined calculation results in an alternative solution algorithm. All authors have read and agreed to the published version of the manuscript.

## Acknowledgements

The authors are grateful to the editor and reviewers for kind guidance and in-depth comments to enable the manuscript to be brushed up.

## Abbreviations

ABM: Agent-Based Model
ATLM: Apparent Time Lag Model (present epidemic model)
DDE: Delay Differential Equation
NPI: Non-Pharmaceutical Intervention
PCR: Polymerase chain reaction
SEIR: Susceptible, Exposed, Infectious and Removed model
SIR: Susceptible, Infectious and Removed model

## Appendix-A

Calculation of the first approximation of M and *α*

In ATLM, the infection rate (number of newly infected persons) 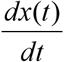 is given by the following equation by the susceptible population M and the infection coefficient *α*.

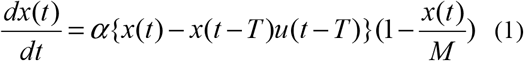

Here, *x*(*t*): cumulative number of infected persons, *T*: number of days from infection to quarantine, u: the step function.

Since the data on the number of newly infected persons varies widely, an integral is used to smooth the data.

The intervals [*t*_1_, *t*_2_],[*t*_3_, *t*_4_](0 < *t*_1_ < *t*_2_ < *t*_3_ < *t*_4_) are used as those of interest.

The following equation is obtained from the integral of (1) in the interval [*t*_1_, *t*_2_].

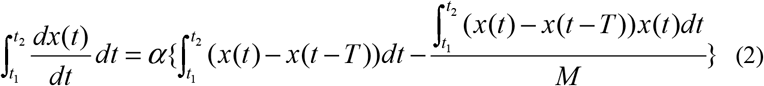

Normally, *T* < *t*_1_. For this reason, we set the step function u = 1.

(2) is modified to obtain (3).

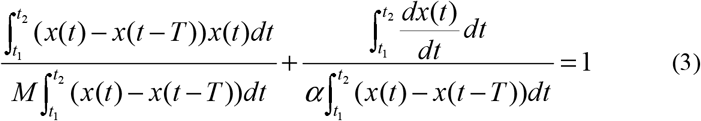

Similarly, (4) is obtained in the interval[*t*_3_, *t*_4_].

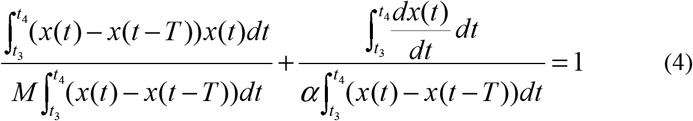

(3) and (4) are linear equations of 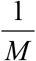 and 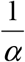. Then M and *α* are calculated.

Usually, the following inequality holds owing to monotony of *x*(*t*).

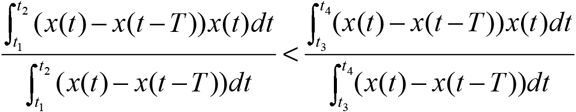

The condition (5) is sufficient for M and *α* to be positive.

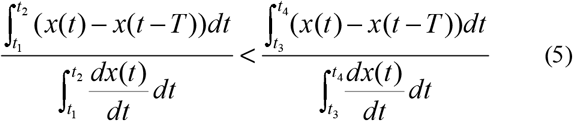

Example of Israel

The source of the number of newly infected persons is as follows.

<u>Israel COVID: 836,926 Cases and 6,334 Deaths - Worldometer (worldometers.info)</u>

The intervals were 2020 11/29 - 12/12 (14days) and 12/13 - 12/26 (14days) before the lockdown. The number of days from infection to quarantine was T = 14 days. Therefore, the measured number of newly infected persons 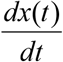 on the day *t* was used as the data of the number of newly infected persons T = 14 days ago.

The cumulative number *x*(*t*) of infected persons was accumulated from the minimum number of newly infected persons on 25 October.

Setting *1/M -> x* and *1/α->y*, we obtained the following two linear equations.

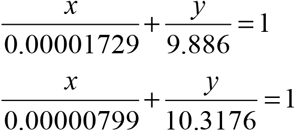

From this equation, M = 1.66714 * 10 ^ 6 and *α* = 0.104788 were obtained. These values are the first approximations.

All integrals were calculated with the trapezoidal formula. Also it was used for the data on the number of newly infected people 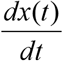. Therefore, numerically, 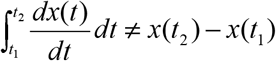 If the number of newly infected persons is accumulated from 11/15, almost the same M and *α* are obtained.

If necessary, these are graphically tuned such that they are consistent with the measurement.

